# Precision Imaging to Evaluate Kaposi Sarcoma (PRIME-KS): protocol for a multicountry novel artificial intelligence-based imaging device

**DOI:** 10.64898/2026.06.03.26354815

**Authors:** Thomas A. Odeny, Harriet Fridah Adhiambo, Dorothy Mangale, Philippa Kadama Makanga, Beryne Odeny, Fred Okuku, Chao Zhou, Elvin H. Geng, Joseph Carson, Victor Mudhune, Elizabeth A. Bukusi, Aggrey Semeere

## Abstract

**Background:** Kaposi sarcoma (KS) is the most common cancer among men in several Eastern African countries, yet treatment monitoring relies on imprecise, time-consuming ruler-based measurements defined by the AIDS Clinical Trial Group (ACTG). This method suffers from inter-observer variability, fails to capture lesion height or true geometric area, and performs poorly on dark skin. SkinScan3D (SS3D) is a portable, low-cost, AI-enabled 3D imaging device that provides objective measurements of KS skin lesion area, height, volume, and color. The Precision Imaging to Evaluate Kaposi Sarcoma (PRIME-KS) study evaluates whether SS3D provides more reproducible and accurate lesion measurements than the standard method, and validates its integration into routine clinical workflows in Kenya and Uganda.

**Methods:** PRIME-KS is a multicountry prospective mixed-methods study with two clinical objectives. Objective 1 is a cross-sectional diagnostic accuracy study comparing SS3D with ruler-based measurement in 50 adults with KS (150 lesions) across sites in Kenya and Uganda. Two clinicians independently measure three lesions per participant using both methods. The primary outcomes are concordance correlation coefficient (CCC) for inter-rater reproducibility, and co-efficient of determination for accuracy. Objective 2 is a non-randomized before-and-after pilot study in 100 patients at three sites, evaluating device usability, acceptability, appropriateness, and feasibility using validated instruments, along with time-and-motion studies and activity-based micro-costing. Prior to these clinical objectives, a formative study used focus group discussions, discrete choice experiments, and human-centered design workshops to refine the SS3D device and protocols with end-user input.

**Discussion:** PRIME-KS will provide the first rigorous evaluation of a 3D imaging device for monitoring KS treatment response in routine clinical settings. If SS3D demonstrates superior reproducibility and clinical utility, it could reduce unnecessary chemotherapy exposure and associated toxicities by enabling earlier, more objective assessment of treatment response.

**Trial registration:** ClinicalTrials.gov NCT06898203, registered 27 March 2025. Pan African Clinical Trials Registry PACTR202603523439856.

**Structured summary:** 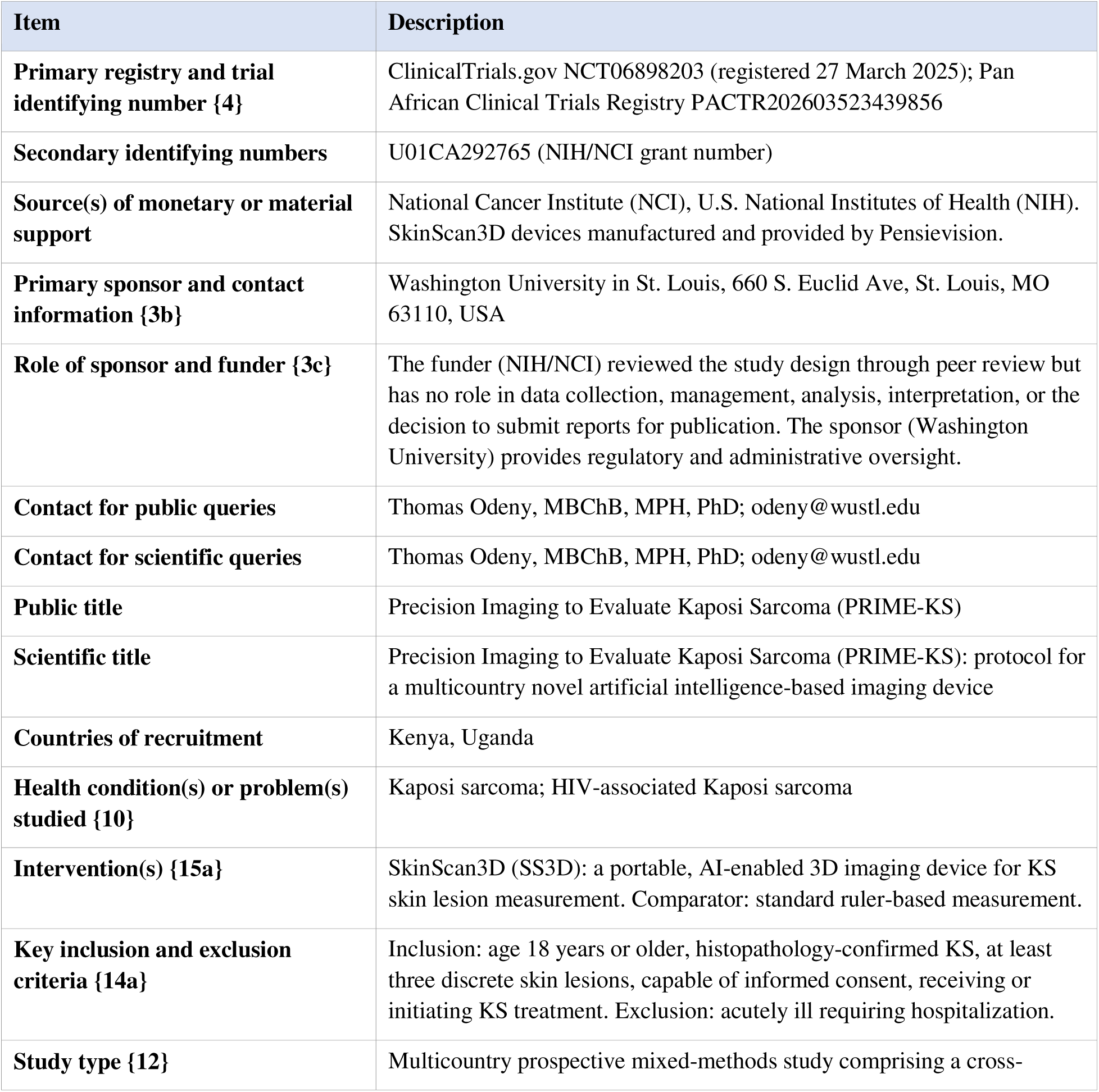

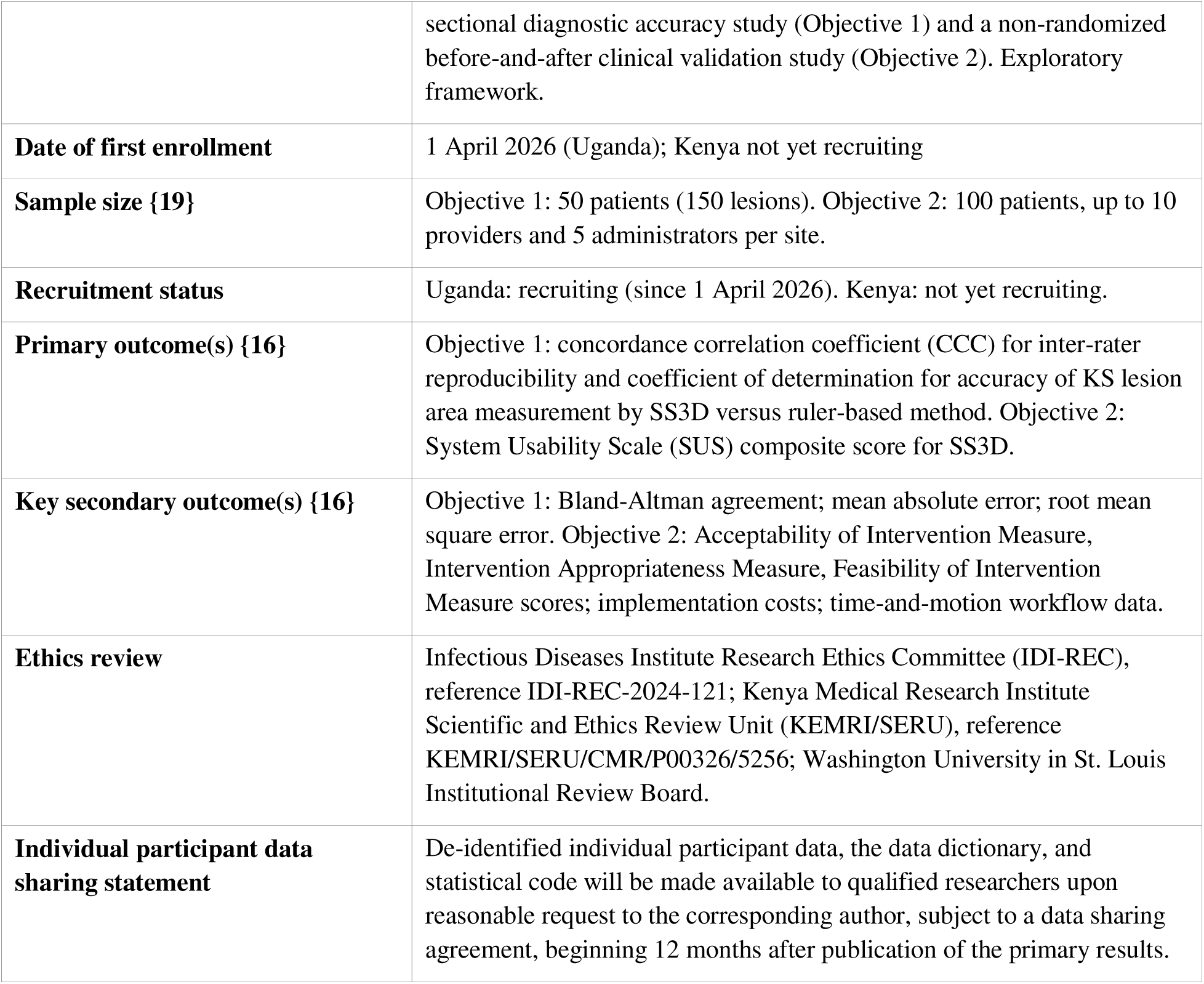

**Protocol version {2}:** Version 1.2, 2 May 2026.

## Introduction

### Background and rationale {9a}

Traditional monitoring of treatment response for Kaposi sarcoma (KS) relies upon user observation and measurement of changes in lesion dimensions based on user observation of color contrast between lesion and normal skin. This may miss subtle changes in lesion pigment that may not be readily apparent, especially in patients with skin of color. In addition, this method suffers from inter-observer variability. Three-dimensional (3D) imaging removes user subjectivity and mitigates problems with identifying and characterizing KS lesion changes on patients by providing an independent measurement of cutaneous tumor volume based on 3D shape with precise calculation of lesion area, instead of the more problematic color contrast and perpendicular lengths.

There is an unmet need to develop simple, accurate, and less burdensome tools for monitoring KS treatment. Currently, response to KS treatment is assessed using manual ruler-based measurements defined by the AIDS Clinical Trial Group (ACTG) Oncology Committee. ^1, 2^ This method involves time-consuming and tedious manual measurement of multiple lesions using a ruler with millimeter precision, leading to inter- and intra-observer variation and failing to capture the true geometric area (Figure 1).

**Figure 1.**
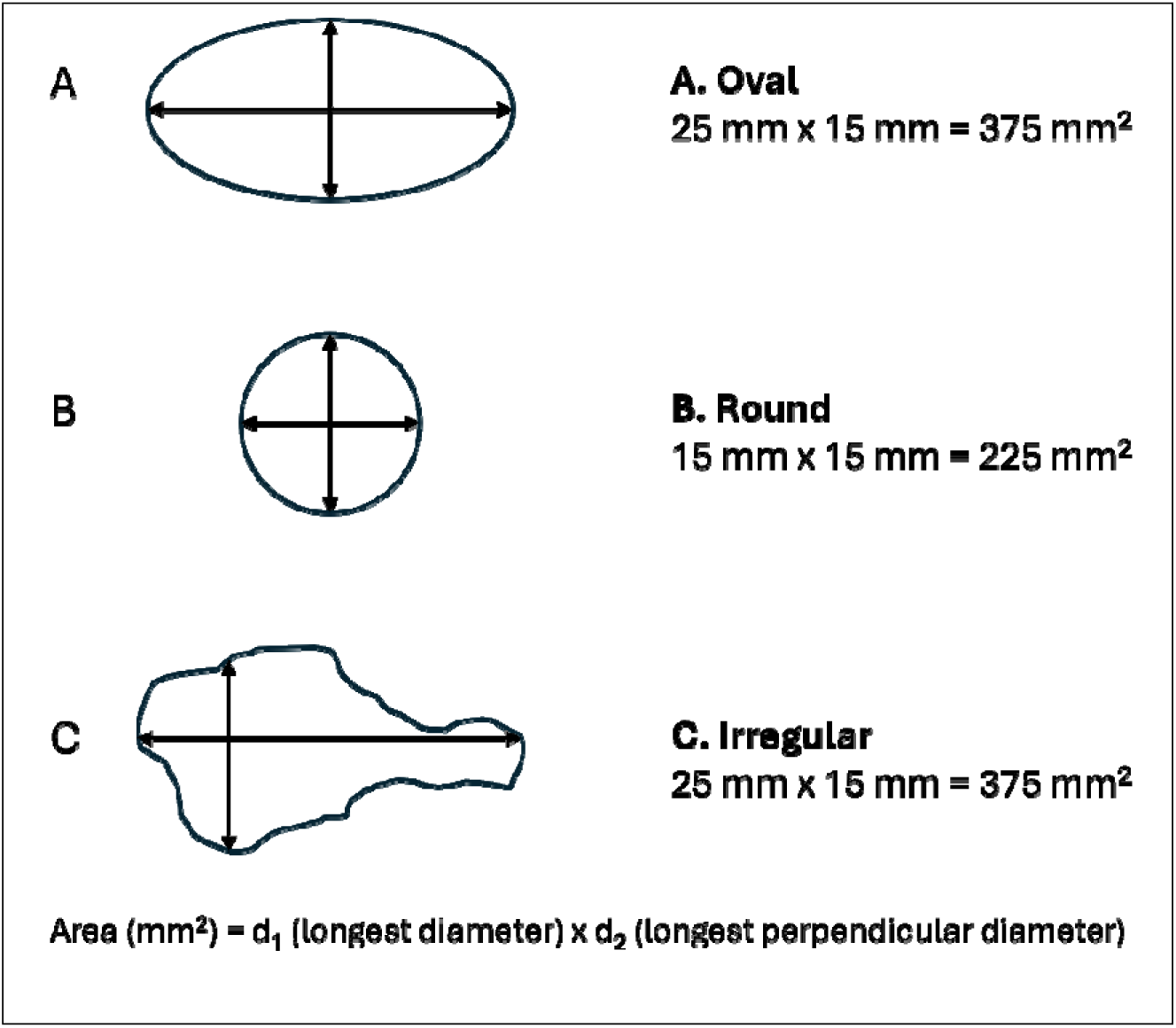
Manual bidimensional measurement for Kaposi sarcoma skin lesions using the AIDS Clinical Trial Group (ACTG) method (not drawn to scale). Three representative morphologies are shown: (A) oval, (B) round, and (C) irregular. Lesion area is calculated as the product of the longest diameter (d_1_) and the longest perpendicular diameter (d_2_), both measured in millimeters using a ruler. This method does not capture lesion height, volume, or color.

The color and degree of nodularity (including lesion height) of KS lesions are difficult to experimentally quantify and are not incorporated into the ACTG method, yet they often provide the most important information in assessing treatment response. ^2^ In settings with high prevalence of KS, the limited number of practitioners trained in evaluating KS treatment response exacerbates the problem. Due to the rigorous training required for competency in this method, it is only readily applicable in highly controlled clinical trial settings. The complex and onerous requirements (e.g. identify bidimensionally measurable KS cutaneous marker lesions, yet lesions appear in multiple shapes and sizes) make it impractical for routine care settings, resulting in possible inaccurate, unreproducible, and suboptimal treatment monitoring. Precise monitoring of KS treatment is essential to facilitate timely treatment adjustments. Treating clinicians primarily rely on subjective observation of treatment response and many err on the side of prolonging treatment with chemotherapy. Introducing a simple and user-friendly device could alleviate the need for extensive training and facilitate its adoption in routine care settings.

The current ACTG criteria primarily rely on reduction in bidimensional tumor size, which is not ideal for the majority of patients who present late with nodular lesions,^3^ that may not exhibit a significant change in bidimensional size. Instead, treatment response in these cases is characterized by a transition from a raised to a flat state and also accompanied by change in color from darker to lighter lesions. This response often precedes absolute reduction in bidimensional lesion size, leading to the unnecessary continuation of cytotoxic chemotherapy and the accumulation of associated toxicities. 3D imaging of lesions could introduce more precise treatment monitoring that is attuned to actual treatment response by detecting these early changes and facilitating early discontinuation of chemotherapy when indicated.

The SkinScan3D (SS3D) imaging device is a hand-held technology capable of measuring KS lesion height, area, and color, crucial determinants of treatment response for nodular lesions.^4^ It is a simple, low-cost, and user-friendly technology that combines liquid lens technology and artificial intelligence (AI), providing high-resolution 3D images/maps of KS skin lesions incorporating lesion height, area, and volume. We will explore the importance of measuring lesion height to obtain previously unmeasured unique clinical properties that may be used to objectively measure efficacy of treatment. We will also explore how changes in lesion color correlate with response assessment. The lack of an effective measurement tool is problematic because, while progress has been made in the last decade in the treatment of KS in Africa and elsewhere, ^5-8^ treatment is not curative and needs simple, reproducible methods to accurately, quantitatively, and reproducibly gauge the response to new treatment modalities.

Various alternative technologies, such as infrared thermal imaging, multi-spectral imaging, ultrasound, radiography, CT, MRI, PET, scintigraphy, and LDI, offer potential for improving tumor measurement. ^9^ However, none of these options provide a practical or affordable solution for clinicians to obtain accurate and reproducible 3D images or maps of cutaneous KS tumors, enabling serial comparisons crucial for assessing treatment response. The pressing clinical need to develop an improved, affordable, simple, accurate, and reproducible tool for monitoring KS treatment response is particularly urgent in East Africa, where KS ranks among the most prevalent cancers.^10^

We propose to evaluate SS3D and its potential contribution in the care of patients with KS by optimizing performance and integrating it into workflows at cancer treatment clinics in Kenya and Uganda. First, we will work with stakeholders to formally refine the technology to make it user-friendly and easy to integrate into clinical workflows. Second, we will compare lesion measurement using SS3D versus the standard manual method. Finally, we will assess SS3D’s contribution to improved clinical outcomes in real-world clinical practice.

### Explanation for the choice of comparator {9b}

The comparator is the current standard of care for KS lesion measurement: manual ruler-based assessment using ACTG criteria, which calculates lesion area as the product of the longest perpendicular diameters (Figure 1).^1, 2^ This method was selected because it is the globally recognized standard for KS treatment response assessment used in clinical trials. Comparing SS3D against this established method will determine whether SS3D offers superior reproducibility and accuracy for clinical decision-making.

### Objectives {10}

**Objective 1:** Compare reproducibility and accuracy of KS lesion size measurements between SkinScan3D and current standard of care measurement.

**Objective 2:** Validate and optimize SkinScan3D clinical workflow in real-world settings.

## Methods: Patient and public involvement, and trial design

### Patient and public involvement {11}

Patient and public involvement is central to PRIME-KS and was formalized through a dedicated formative study that preceded and informed the clinical objectives described in this protocol.

This formative work engaged patients with KS, healthcare providers, and community representatives in refining the SS3D device and usage protocols prior to clinical evaluation.

The formative study used a sequential mixed-methods approach comprising three components. First, focus group discussions (FGDs) were conducted with three stakeholder groups (patients with KS, end-users such as oncologists, dermatologists, clinical officers, and nurses, and community representatives from patient advocacy groups) in both Kenya and Uganda. In-depth interviews (IDIs) were conducted with up to 15 additional community stakeholders including community advisory board members, patient advocates, caregivers, and medical regulatory authorities. These qualitative activities were designed to understand current perspectives on KS skin lesion assessment, identify barriers and facilitators to adopting new technologies, and assess local cultural sensitivities and preferences that may influence acceptability and adoption of SS3D.

Second, findings from the FGDs and IDIs informed the development of a discrete choice experiment (DCE) survey that quantified end-user preferences for various design features of SS3D (e.g., camera resolution, data processing time, accessibility, ease of use, language support).

Third, iterative human-centered design (HCD) workshops were conducted using the Discover, Design/Build, and Test (DDBT) framework. These co-creation workshops brought together previous FGD and DCE participants, research team members, and technology experts for structured prototyping sessions. Participants were trained on SS3D use and asked to verbalize their experiences using a think-aloud protocol. Design activities yielded visualized outputs (drawings, pictures, paper prototypes, participant-written insight statements) and ethnographic field notes. Results were submitted to the manufacturer (Pensievision) for device redesign of hardware, software, and user guides, while local research teams refined standard operating procedures.

From this formative work, we synthesized findings and developed a refined SkinScan3D package with optimized hardware, software, and SOPs, while maintaining already present specific desirable attributes (operation without electricity or internet, no moving parts for durability, internal self-checks, self-calibration). The outputs of this formative work directly shape the SS3D intervention package evaluated in the clinical objectives of this protocol.

### Trial design {12}

PRIME-KS uses a two-objective sequential design within an exploratory framework. Objective 1 is a cross-sectional diagnostic reproducibility and accuracy study comparing SS3D with ruler-based KS lesion measurement. It uses a paired design in which two clinicians independently measure three lesions per participant using both methods. Objective 2 is a non-randomized before-and-after pilot study in which providers first use the manual ruler-based method for three months, then switch to using SS3D, allowing within-provider comparison of implementation outcomes. There is no randomization; the study is a prospective single-arm observational study to validate accuracy, reproducibility, and clinical workflow integration.

## Methods: Participants, interventions and outcomes

### Trial setting {13}

The Kenya sites are Homa Bay County Referral Hospital and the Kenya Medical Research Institute (KEMRI) located within the Jaramogi Oginga Odinga Teaching and Referral Hospital (JOOTRH) compound in Kisumu, Kenya. JOOTRH is the largest cancer treatment center and tertiary referral hospital in western Kenya, serving a population of over 10 million people.

KEMRI houses the NIH Division of AIDS Clinical Trial Unit and has hosted major KS treatment trials including AMC 066/ACTG A5263. Uganda site activities will be implemented at the Infectious Diseases Institute (IDI) HIV clinic and the Uganda Cancer Institute (UCI) located within the Mulago Hospital Complex in Kampala, Uganda. UCI has been the national cancer treatment, research, and teaching center since 1967 and has hosted multiple KS studies. IDI provides coordination support and patient referral for KS diagnosis.

### Characteristics of the people who are needed for the trial

This study will enroll all adults aged 18 years or older diagnosed and treated for KS who consent to participate in Kenya and Uganda. Objective 1 will include adult patients with a confirmed diagnosis of KS eligible for chemotherapy. In Objective 2, we will include both adult patients with KS undergoing treatment, providers (oncologists, medical officers, and nurses), and administrators involved in delivering KS treatment.

### Eligibility criteria for participants {14a}

#### Objective 1 (Reproducibility and Accuracy)

**Inclusion criteria:** Age 18 years or older; histopathology-based KS diagnosis; at least three discrete skin lesions; capable of informed consent; on treatment for KS (chemotherapy and/or antiretroviral therapy).

**Exclusion criteria:** Patients not initiating KS treatment or very ill and requiring hospitalization.

#### Objective 2 (Clinical Validation)

**Patient inclusion criteria:** Histopathology-based KS diagnosis; at least three discrete skin lesions; age 18 years or older; capable of informed consent; initiating treatment for KS (chemotherapy and/or antiretroviral therapy).

**Patient exclusion criteria:** Patients, not willing to participate.

**Provider and administrator inclusion criteria:** Oncologists, medical officers, nurses, and administrators involved in delivering KS treatment at a participating site.

### Eligibility criteria for sites and those delivering interventions {14b}

The pre-selected participating sites are health facilities that provide routine KS treatment services in Kenya and Uganda, with institutional support for study implementation and capacity for ethical review and regulatory compliance. Clinicians delivering the SS3D intervention at each site will complete a structured training program that includes didactic instruction on the ACTG measurement criteria and practical sessions on SS3D device operation and the standardized measurement protocol.

### Who will take informed consent? {32a}

Informed consent from potential trial participants will be obtained by trained research assistants. Study staff will provide potential study participants with sufficient information about the study, details of the data collection component they will participate in, and their role in the study. This includes a brief video describing the device and its function that will be shared with participants during the consenting process. All consent forms will be translated into the local languages (Kiswahili, Dholuo, Luganda) and back-translated into English to ensure correct use of language.

Health facility and study staff will be on hand to answer any questions that arise before signing the form. We anticipate that a minority of eligible patients may not be able to read or write. In this case, study staff will read the consent document to the eligible participant who will then be asked to place a thumbprint on the signature line. Reading and signing of consent documents will be witnessed and the witness will sign the informed consent form.

### Additional consent provisions for collection and use of participant data and biological specimens {32b}

No biological specimens are collected in this study. The informed consent form includes provisions for the collection and storage of de-identified imaging data (3D images of skin lesions) for the purposes of device calibration, algorithm improvement, and future research. Participants may decline permission for future use of imaging data while still participating in the study. If future scientific aims require biological specimens, the study protocol will be amended in line with institutional ethical review procedures at each site.

### Intervention and comparator

#### Intervention and comparator description {15a}

**Intervention (SkinScan3D):** SS3D is a portable, handheld 3D imaging device manufactured by Pensievision. It uses a photon recording device combined with active optics capable of rapid change of focus, thereby achieving one-click, all-focus, 3D imaging using only a single perspective. The device consists of a custom-designed light ring (enabling white- and colored-light imaging), an electronically controlled liquid lens (Corning Varioptic A-16F0), an 8-megapixel camera, and a Raspberry Pi 4 Model B microcomputer with a 6-inch touchscreen display, all housed in a polycarbonate enclosure. A cup-shaped front-end adaptor ensures fixed shooting angle, camera-to-skin distance, and controlled illumination. A 73-frame image set requiring 8-9 seconds to collect is processed by AI software to generate a 3D map. The device provides automated measurements of lesion area, height, volume, and color. It operates on an internal lithium battery without requiring electricity or internet connectivity. The disposable front-end adaptor is the only consumable component. For each participant, five 3D snapshots of each of three target lesions are collected.

**Comparator (ruler-based measurement):** Participants will have three discrete skin lesions measured using the standard ACTG method, in which lesion area is determined as the product of the longest perpendicular diameters using a ruler with millimeter precision (Figure 1).^1, 2^ This is the current global standard for KS response assessment in clinical trials.

#### Criteria for discontinuing or modifying allocated intervention/comparator {15b}

There are no anticipated criteria for modifying the allocated measurement method during the study. Participants may withdraw from the study at any time.

#### Strategies to improve adherence to intervention/comparator {15c}

All clinicians will receive structured training on both measurement methods prior to data collection, including didactic instruction and supervised practice sessions. A standardized measurement protocol and clinical checklist will guide device operations at each encounter. The SS3D software includes built-in self-checks that alert the user to hardware errors and data quality issues. Regular site monitoring visits and study team meetings will reinforce protocol adherence.

#### Concomitant care permitted or prohibited during the trial {15d}

All participants will continue to receive their standard KS treatment (chemotherapy and/or antiretroviral therapy) as directed by their treating clinicians. The study does not modify or restrict any aspect of clinical care. SS3D measurements are performed in addition to, not instead of, standard clinical assessments during the study period.

#### Ancillary and post-trial care {34}

All participants will continue to receive standard KS care at their treating institution regardless of study participation. There is no additional post-trial care obligation, as the study intervention (3D imaging) poses minimal risk. In the unlikely event that a participant suffers harm directly attributable to study participation, appropriate medical care will be provided at the study site at no cost to the participant.

### Outcomes {16}

#### Objective 1 (Reproducibility and Accuracy)

**Primary outcomes:** (1) Concordance correlation coefficient (CCC) for inter-rater reproducibility of KS lesion area measurement. The CCC will be calculated separately for SS3D and for the ruler-based method, based on measurements by two independent clinicians of the same three lesions per participant. The measurement variable is projected lesion area (mm^2^). The method with the higher CCC will be considered the more reproducible. (2) Coefficient of determination (R-squared) for each measurement method, assessing the proportion of variation attributable to true lesion size versus random measurement error.

**Secondary outcomes:** (1) Bland-Altman agreement plots comparing the two methods. (2) Mean absolute error and root mean square error. (3) Exploratory analyses of correlation between area measurements and changes in lesion height and color, stratified by lesion morphology (flat, nodular, plaque) and size (less than 1 cm versus 1 cm or greater).

#### Objective 2 (Clinical Validation)

**Primary outcome:** System Usability Scale (SUS) composite score for SS3D, measured after device use. The SUS is a validated 10-item questionnaire yielding a single score from 0 to 100, with scores above 68 considered above average.^11^

**Secondary outcomes:** (1) Acceptability of Intervention Measure (AIM), Intervention Appropriateness Measure (IAM), and Feasibility of Intervention Measure (FIM) scores.^12^ (2) Time-and-motion data characterizing clinic workflows, task-shifting, and time associated with clinic visits. (3) Implementation cost per patient and per visit, estimated using activity-based micro-costing.^13^

### Harms {17}

The risks posed to participants are minimal, as the study involves non-invasive imaging and questionnaire completion. There is a very low risk of loss of privacy if there was a breach in data security. We have not experienced such a breach with our current systems that feature limited access to personnel and password-protected authentication among approved users. Patients may also experience emotional discomfort when talking about their skin conditions. To mitigate, we will train our interviewers to observe participants for discomfort, minimize probing when discomfort is perceived, and express empathy to support the participant. All adverse events observed or reported will be documented and reported to the relevant ethics committees according to their requirements.

### Participant timeline {18}

#### Objective 1

Each participant will complete a single study visit. At this visit, written informed consent will be obtained, demographic and clinical data collected, and three marker lesions identified. Two clinicians will independently measure the three lesions using both the ruler-based method and SS3D. The total additional time for study procedures is estimated at 15 to 30 minutes beyond the routine clinic visit.

#### Objective 2

During months 1 to 3 (control period), providers will measure KS lesions using the standard ruler-based method. Providers will also complete training on SS3D during this period.

Thereafter, self-administered survey questionnaires will be used to gather information on usability, acceptability, appropriateness, and feasibility of the SS3D intervention. During the intervention period, providers will switch to using SS3D for KS lesion measurements. Time-and-motion observations and cost data collection will occur throughout both periods. Semi-structured interviews with a purposive sample of providers will then be conducted after the intervention period.

### Sample size {19}

Objective 1: Assuming that the mean and variance of the area of lesions (as determined by the SkinScan3D method) are the same for two clinicians, then the expected width of a 95% confidence interval (CI) for the concordance correlation coefficient for a given number of lesions examined is shown in Table 1. Therefore, with at least 50 participants with 3 lesions each (150 lesions), we will get a precise estimate of the CCC. To address overinterpretation of the concordance, we will complement the R-squared with additional metrics that capture accuracy more directly, including Bland-Altman plots to assess agreement and systematic bias, mean absolute error, and root mean square error. These measures will allow us to evaluate both correlation and the magnitude and direction of differences between methods.

**Table 1.**
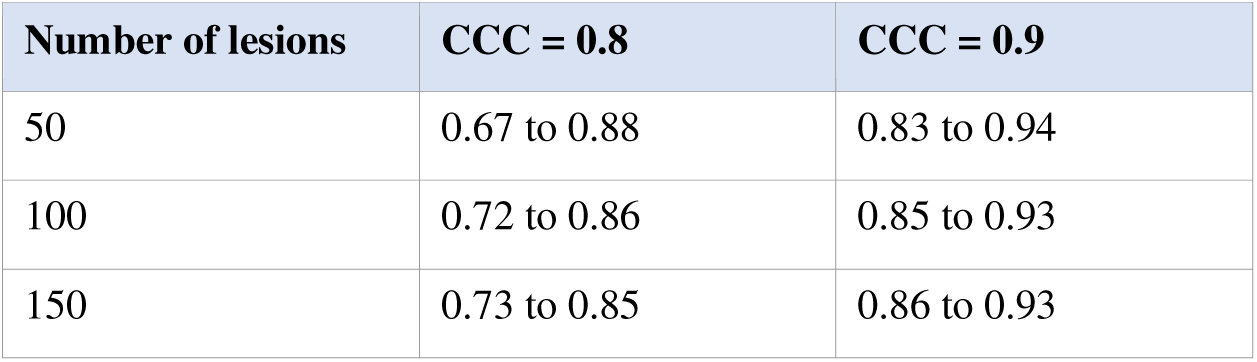
Expected width of 95% confidence intervals (precision)

| Number of lesions | CCC = 0.8 | CCC = 0.9 |
| --- | --- | --- |
| 50 | 0.67 to 0.88 | 0.83 to 0.94 |
| 100 | 0.72 to 0.86 | 0.85 to 0.93 |
| 150 | 0.73 to 0.85 | 0.86 to 0.93 |

Objective 2: We will roll out the refined SS3D intervention to 100 participants (50 patients at UCI; 25 at JOOTRH; 25 at Homa Bay). All providers will complete quantitative surveys. We anticipate recruiting providers (up to n=10), clinic administrators and key decision makers (up to n=5) who will participate in semi-structured interviews at each clinic. The final sample of semi-structured interviews will be determined by the point at which thematic saturation is achieved.

### Recruitment {20}

Objective 1: Eligible patients will be identified from all new patients referred for KS treatment at each site. Recruitment will occur during waiting times to minimize disruption to patient flow. At the screening visit, staff will obtain written informed consent including discussion of the voluntary nature of research, the specific goals of the study, study procedures, and the potential risks and benefits of study participation.

Objective 2: At each site, providers involved in KS treatment will be requested to participate. They will be recruited by cadre to ensure representation of all providers and administrative roles for quantitative and qualitative evaluations. Eligible patients attending clinic visits will be sequentially sampled to participate in both the clinical evaluation and time-and-motion studies.

### Assignment of interventions: Not applicable

Sequence generation: {21a} Not applicable. This is a non-randomized observational study. Sequence generation: {21b} Not applicable.

Allocation concealment mechanism {22} Not applicable. Implementation {23} Not applicable.

### Assignment of interventions: blinding

Who will be blinded {24a} Not applicable.

How will blinding be achieved {24b} Not applicable. Procedure for unblinding if needed {24c} Not applicable.

### Data collection and management

Plans for assessment and collection of outcomes {25a}

#### Objective 1 measurements

For each participant we will collect demographic and clinical data regarding KS diagnosis and extent of disease. Thereafter, two clinicians will each measure three lesions using both the manual ruler-based method (standard of care) and SS3D. Standard of care measurement: Participants will have three discrete skin lesions measured, with lesion area determined by ACTG criteria utilizing the product of longest perpendicular diameters. Participants will also have five 3D snapshots of each of the three target lesions taken, measuring lesion area automatically using SS3D AI.

#### Objective 2 measurements

Measurements for Objective 2 will happen in two phases. In phase 1, providers will first measure KS lesions using the manual ruler-based method as per usual standard of care for the first three months. During this time, providers will be trained on use of the adapted SS3D device and protocols. Thereafter, we will administered survey questionnaires to gather information on usability, acceptability, appropriateness, and feasibility of the SS3D intervention. We will also conduct direct observation for workflow mapping, time-and-motion studies, and micro-costing within implementing clinics. Using the System Usability Scale (SUS)^11^, a reliable, well-validated ten-item scale for measuring usability of hardware, software, and applications, we will collect information from providers on multiple aspects of system usability, including need for support, training, complexity. Data regarding acceptability, appropriateness, and feasibility of integrating the device into routine workflows will be collected using the following validated instruments: the Acceptability of Intervention Measure (AIM), Intervention Appropriateness Measure (IAM), and Feasibility of Intervention Measure (FIM)^12^. We will collect data on clinic workflows including any task-shifting, and time associated with clinic visits for patients and providers through time-and-motion studies. Implementation costs will be documented using activity-based micro-costing techniques ^13^.

We will select a random sample of providers to undergo semi-structured interviews to enrich findings from the quantitative measures above.

### Plans to promote participant retention and complete follow-up {25b}

Participants are patients receiving ongoing KS treatment at the study sites and are seen as part of routine clinical care. Retention is promoted through alignment of study activities with routine clinic visits. Patients who discontinue KS treatment or transfer care will have the reason documented.

### Data management {26}

All structured quantitative data will be stored in a centralized, web-based data management scheme using REDCap.^14^ Password-protected access will be strictly limited to study investigators and personnel. For the qualitative data, all audio recordings, transcripts, and results from coding will be stored in password-protected computers. Qualitative data will be recorded as audio files, transcribed, and coded using Dedoose.^15^

### Confidentiality {33}

All raw and analysis data will be stored within a secure computing environment. All potential identifiers will be removed prior to data analysis. Participants will be assigned a unique study identification number at enrollment. All data will be recorded and stored using this identifier rather than personal information. No participant-identifying information will be disclosed in any report or publication arising from the study. SS3D images of skin lesions are stored with the study identification number only. Physical documents containing identifying information are stored in locked cabinets at each site with access restricted to the site principal investigator and designated study staff. Electronic data are protected by encryption and role-based access controls.

## Statistical methods

Statistical methods for primary and secondary outcomes {27a}

### Objective 1

We will plot a graph for each measurement method. Each graph’s x-axis will represent measurements from clinician 1 and y-axis will represent measurements from clinician 2. For each graph, we will calculate the concordance correlation coefficient (CCC) and coefficient of determination (R-squared). The CCC measures reproducibility (agreement) between clinicians for each method. The method with the higher CCC will be considered the more reproducible, and the method with R-squared closer to 1 will be considered the more accurate. We will also visualize agreement between methods graphically using Bland-Altman plots, and report the mean difference and 95% limits of agreement. To provide a more robust assessment, we will also report mean absolute error and root mean square error to capture the magnitude of differences between methods.

### Objective 2

System Usability Scale (SUS) scores will be computed following standard scoring procedures (score contributions from items summed and multiplied by 2.5, yielding a score range of 0 to 100). Device usability will further be normalized to generate percentile ranks and graded as either acceptable (score above 68), marginal, or not acceptable. AIM, IAM, and FIM scores will be presented as means, standard deviations, and ranges. Qualitative data collected from semi-structured interviews will be transcribed, coded, and thematically analyzed to identify sub-themes and themes related to SkinScan3D implementation in routine clinical settings. Data triangulation will be applied to cross-validate findings from interviews and direct observation to ensure rigor and validity of the qualitative analysis.

For the cost analysis, we will summarize data to estimate the total program cost of SS3D per patient and per visit, excluding research costs. Costs will be broken down by the categories in Table 2 to identify key cost drivers.

**Table 2.** Summary of cost measures and associated data sources.

| Cost type | Elements | Data sources |
| --- | --- | --- |
| **Start-up** | Training materials; training expenses (trainer fees, staff time spent in training) | Project expense reports; staff/civil service salaries |
| **Implementation** | Training expenses; technical assistance; fidelity monitoring; community outreach | Project expense reports |
| **Personnel** | Time/salaries of healthcare personnel, supervisors, and administrative staff | Direct observations; salaries; interviews |
| **Patient** | Transportation; other out-of-pocket expenses including childcare; time losses | Patient surveys; local average wages |
| **Supplies** | Consumables utilized or dispensed on a per-patient basis, particularly front-end adaptors | Clinic and project expense reports |
| **Capital** | SkinScan3D equipment utilized in clinics | Manufacturing and shipping costs |
| **Utilities** | Space/storage, internet, electricity, other overhead costs associated with SS3D | Clinic and project expense reports |

### Who will be included in each analysis {27b}

All enrolled patients with complete paired measurements will be included. Analysis for Objective 1 will be at the lesion level (up to 150 lesions from 50 participants). For Objective 2, the usability and implementation outcome analyses will include all providers who completed the relevant questionnaires. The cost analysis will include all patients enrolled during the study period.

### How missing data will be handled in the analysis {27c}

For Objective 1, if a measurement cannot be completed for a particular lesion using one method (e.g., SS3D cannot image a lesion on a highly curved surface), that lesion will be excluded from the paired analysis for that method. We will report the proportion and reasons for missing measurements. Given the cross-sectional, single-visit design, we anticipate minimal missing data. No imputation is planned for Objective 1. For Objective 2, if a provider does not complete a questionnaire, available data will be analyzed and the extent of missing data reported. For the cost analysis, missing cost elements will be estimated using median values from the available data at the same site.

Methods for additional analyses (e.g. subgroup analyses) {27d}

For Objective 1, exploratory subgroup analyses will examine CCC and R-squared stratified by lesion morphology (flat versus nodular versus plaque), lesion size (less than 1 cm versus 1 cm or greater), and study site. We will also explore the correlation of area measurements with changes in lesion height and color. For Objective 2, implementation outcomes will be described by site and by provider cadre.

### Interim analyses {28b}

No formal interim analyses or stopping rules are planned. This is an exploratory study evaluating a diagnostic device, and there is no clinical equipoise that would necessitate early stopping. The study team will review enrollment and data quality on an ongoing basis through regular study team meetings.

### Protocol and statistical analysis plan {5}

The full study protocol is available through this publication. The statistical analysis plan is integrated into this protocol (see sections {27a} through {27d}).

### Oversight and monitoring

Composition of the coordinating centre and trial steering committee {3d}

The study is coordinated by Washington University School of Medicine (Division of Medical Oncology), which serves as the central coordinating site responsible for overall study management, regulatory compliance, data management, and biostatistical analysis. The Kenya site is coordinated by KEMRI. The Uganda site is coordinated by the Infectious Diseases Institute in collaboration with the Uganda Cancer Institute. A study executive committee comprising the principal investigators (TAO, AS), the device engineer (JC), and site coordinators meets regularly by teleconference to review study progress, enrollment, data quality, and operational issues.

Composition of the data monitoring committee, its role and reporting structure {28a}

A formal data monitoring committee (DMC) was not established for this study. This decision is justified by the minimal risk nature of the intervention (non-invasive imaging of skin lesions), the absence of a therapeutic intervention that could cause harm, and the exploratory rather than confirmatory design of the study. Data quality and study conduct are monitored by the study executive committee and through regular site monitoring visits.

Frequency and plans for auditing trial conduct {29}

The coordinating center will conduct remote monitoring of data quality monthly through REDCap data reports, including enrollment tracking, data completeness, and range check violations. On-site monitoring visits will be conducted at each site at least twice during the study period (at approximately 25% and 75% of target enrollment) to verify source data, review consent documentation, assess protocol adherence, and address operational issues. Additional visits may be triggered by identified data quality concerns.

Protocol amendments {31}

Any substantive amendments to the protocol will be submitted to the relevant ethics review committees (IDI-REC in Uganda; KEMRI SERU in Kenya; Washington University IRB in the United States) for review and approval prior to implementation. Amendments will also be updated in the ClinicalTrials.gov registry (NCT06898203) and the Pan African Clinical Trials Registry (PACTR202603523439856). All investigators will be notified of approved amendments. Administrative or non-substantive changes will be documented in an amendment log and communicated to the ethics committees at the next scheduled review.

Dissemination policy {8}

Results will be disseminated through multiple channels. Primary findings will be submitted for publication in peer-reviewed journals. Abstracts will be submitted to scientific conferences.

Results will be reported in the ClinicalTrials.gov and PACTR registries. Plain-language summaries will be shared with community advisory boards and patient advocacy groups at each site. Authorship on future trial publications will follow ICMJE guidelines.

## Discussion

PRIME-KS will provide the first rigorous evaluation of a 3D imaging device for monitoring KS treatment response in routine clinical settings. The study addresses a critical gap in KS care: the absence of a simple, objective, and affordable tool for treatment monitoring that can function effectively, including in resource-constrained settings.

This study provides a unique participatory approach to device development from all stakeholders, ensuring that the SS3D device and protocols are contextually appropriate, acceptable, and designed with the input of the people who will use and benefit from the technology. The formative study activities described under Patient and Public Involvement directly shaped the intervention prior to clinical evaluation, providing a blueprint for participatory approaches to medical device design and implementation in low- and middle-income countries.

Several design considerations merit discussion. The paired within-participant design of Objective 1, in which two clinicians independently measure the same lesions using both methods, is a strength that allows direct comparison of reproducibility while controlling for patient-level variation. The before-and-after design of Objective 2 was chosen over a randomized design because the goal is to characterize the implementation profile of SS3D (usability, acceptability, cost) rather than to demonstrate efficacy; a non-randomized design is appropriate for this purpose and is more feasible given the cluster-level nature of the intervention.

Potential limitations include the lack of a true gold standard for KS lesion measurement. We compare SS3D against the current standard of care rather than against a reference standard such as optical coherence tomography, which is impractical in field settings. Another limitation is that the before-and-after design of Objective 2 is subject to temporal confounding; however, our primary interest is in the implementation profile of the device, for which this design is appropriate. There are also potential challenges of users not fully understanding the technology since it is new, which may yield inappropriate responses during initial implementation; the formative co-creation with potential users, structured training protocol, and think-aloud usability testing are designed to mitigate this.

If SS3D demonstrates superior reproducibility, accuracy, and clinical utility, it could transform KS treatment monitoring by enabling earlier detection of treatment response, informing timely discontinuation or switch of chemotherapy, decreasing the burden on providers in busy clinics, and introducing a standardized measurement method in KS clinical trials and routine care settings.

## Trial status

Protocol version 1.2, dated 2 May 2026. Uganda recruitment began on 1 April 2026. Kenya has not yet begun recruiting. Recruitment is anticipated to be completed by Q4 2027. The formative study activities (Patient and Public Involvement) have been completed.

## Data Availability

No datasets were generated or analyzed for this protocol manuscript. Data generated during the conduct of the study will be made available upon reasonable request to the corresponding author, subject to a data sharing agreement.

## Abbreviations

ACTG: AIDS Clinical Trial Group
AI: artificial intelligence
AIM: Acceptability of Intervention Measure
CCC: concordance correlation coefficient
CT: computed tomography
DCE: discrete choice experiment
DDBT: Discover, Design/Build, and Test
FGD: focus group discussion
FIM: Feasibility of Intervention Measure
HCD: human-centered design
HIV: human immunodeficiency virus
IAM: Intervention Appropriateness Measure
IDI: Infectious Diseases Institute
IDI-REC: Infectious Diseases Institute Research Ethics Committee
JOOTRH: Jaramogi Oginga Odinga Teaching and Referral Hospital
KEMRI: Kenya Medical Research Institute
KS: Kaposi sarcoma
LDI: laser Doppler imaging
MAE: mean absolute error
MRI: magnetic resonance imaging
PET: positron emission tomography
RMSE: root mean square error
SERU: Scientific and Ethics Review Unit
SOP: standard operating procedure
SS3D: SkinScan3D
SUS: System Usability Scale
UCI: Uganda Cancer Institute

## Declarations

## Acknowledgements

The authors acknowledge the patients, healthcare providers, and community members who participate in this study. We thank Susan Krown, MD, for her advisory role on staging and response criteria for KS. Joseph Carson’s contributions were supported in part by U.S. National Science Foundation Award 2242812 and U.S. National Institutes of Health Award P20GM103499.

## Authors’ contributions {3a}

TAO is the principal investigator; he conceived the study, led the proposal and protocol development, and oversees study implementation. AS co-led proposal and protocol development and leads study implementation in Uganda. JC designed and manufactured the SkinScan3D device and leads device refinement and calibration. CZ leads cross-validation and phantom studies for SS3D. BO designed and supervised the human-centered design workshops and qualitative data collection. EHG provided expertise in implementation science and study design. HFA coordinates data collection and analysis at the Kenya sites. DM leads time-and-motion and micro-costing data collection and analysis. FO trains clinicians on lesion measurement and oversees clinical procedures at UCI. PK coordinates study activities in Uganda. VM oversees local regulatory approvals and implementation in Kenya. EB provided expertise in research ethics, collaborative management, and study implementation in Kenya. All authors read and approved the final manuscript. Authorship on future trial publications will be determined according to ICMJE guidelines.

## Sources of funding and other support {7a}

This study is funded by the National Cancer Institute (NCI), U.S. National Institutes of Health, grant number U01CA292765. The SkinScan3D devices are manufactured and provided by Pensievision. The funders had no role in the design of the study, the writing of this protocol, or the decision to submit for publication. The funders will have no role in data collection, analysis, interpretation, or the decision to publish trial results.

## Availability of data and materials {6}

The final trial dataset will be accessible to the principal investigators (TAO, AS) and co-investigators. De-identified individual participant data, the data dictionary, and statistical code will be made available to qualified researchers upon reasonable request to the corresponding author, subject to a data sharing agreement, beginning 12 months after publication of the primary results. There are no contractual agreements that limit investigator access to the data.

## Ethics approval and consent to participate {30}

This study has been approved by the Infectious Diseases Institute Research Ethics Committee (IDI-REC), reference number IDI-REC-2024-121; the Kenya Medical Research Institute Scientific and Ethics Review Unit (KEMRI/SERU), reference number KEMRI/SERU/CMR/P00326/5256; and the Washington University in St. Louis Institutional Review Board. Written, informed consent to participate will be obtained from all participants.

## Consent for publication

Not applicable. No individual person’s data in identifiable form are presented in this protocol.

## Competing interests {7b}

The authors declare that they have no competing interests.

## Notes

### Competing Interest Statement

The authors have declared no competing interest.

### Clinical Trial

NCT06898203

### Author Declarations

This study involves human participants and has received ethical approval from the Washington University in St. Louis Institutional Review Board, the Kenya Medical Research Institute Scientific and Ethics Review Unit (KEMRI/SERU), and the Infectious Diseases Institute Research Ethics Committee (IDI-REC). Written informed consent is obtained from all participants.

